# Estimation of preventable pneumococcal disease burden by PCV20 or PCV21 under the national immunization program for adults aged 65 years in Japan

**DOI:** 10.64898/2026.09.15.26363097

**Authors:** Kosuke Tamura, Bin Chang, Reiko Shimbashi, Kenji Gotoh, Yoshinari Tanabe, Koji Kuronuma, Kengo Oshima, Takaya Maruyama, Msashi Nakamatsu, Shuichi Abe, Kei Kasahara, Tadasuke Ooka, Yu Arakawa, Yuki Kinjo, Yukihiro Akeda, Kazunori Oishi, the Adult IPD Study Group

**Affiliations:** Department of Research Planning, Toyama Institute of Health, Toyama, Japan; Department of Bacteriology I, National Institute of Infectious Diseases, Tokyo, Japan; Department of Infectious Disease Surveillance, National Institute of Infectious Diseases, Tokyo, Japan; Department of Infection Control and Prevention, Kurume University School of Medicine, Fukuoka, Japan; Department of Respiratory Medicine, Niigata Prefectural Shibata Hospital, Niigata, Japan; Department of Respiratory Medicine and Allergology, Sapporo Medical University School of Medicine, Hokkaido, Japan; Department of Infectious Disease, Tohoku University Graduate School of Medicine, Miyagi, Japan; Mie Prefectural Ichishi Hospital, Mie, Japan; Department of Infectious, Respiratory and Digestive Medicine, Graduate School of Medicine, University of the Ryukyus, Okinawa, Japan; Department of Infection Control and Prevention, Yamagata Prefectural Central Hospital, Yamagata, Japan; Center for Infectious Diseases, Nara Medical University, Nara, Japan; Department of Microbiology, Kagoshima University Graduate School of Medical and Dental Sciences, Kagoshima, Japan; Department of Clinical Infectious Diseases, Kochi Medical School, Kochi University, Kochi, Japan; Department of Bacteriology, The Jikei University School of Medicine, Tokyo, Japan; Department of Bacteriology, Toyama Institute of Health, Toyama, Japan

**Author notes:** **Correspondence:** Kazunori Oshi, M.D., Ph.D. 17-1 Nakataikouyama, Imizu, Toyama Prefecture, Japan.

**Keywords:** PCV20, PCV21, IPD, adults, Japan, pneumococcal community-acquired pneumonia Word count, 3, 488 words

## Abstract

**Aims:** To estimate the preventable pneumococcal disease burden by 21-valent pneumococcal conjugate vaccine (PCV21) compared to PCV20 for the national immunization program for adults aged 65 years or older in Japan.

**Methods:** Serotype distribution among isolates and the annual incidence of invasive pneumococcal disease (IPD) in adults were monitored. The numbers of IPD or pneumococcal community-acquired pneumonia (pCAP) cases preventable by immunization with PCV20 or PCV21 were estimated according to reported vaccine efficacy and duration of protection in older adults. Data from our nationwide study of adult IPD surveillance and a multicenter study of pCAP were used.

**Results:** The mean annual incidence of adult patients with IPD caused by all serotypes among adults aged ≥65 years, which declined during the COVID-19 pandemic period, recovered to 89% of the pre-pandemic level in 2025. During 2023–2025, the proportion of PCV20-non-PCV21 serotypes was 8.5% for adults aged ≥65 years. At the current vaccination coverage rate of 40%, the numbers of preventable IPD cases (per person per year) by PCV20 and PCV21 were 36 and 53, respectively. The preventable numbers of pCAP cases by PCV20 and PCV21 were 2,821 and 4,765, respectively. The estimated number of preventable pCAP cases was approximately 80–90 times higher than that of preventable IPD cases.

**Conclusion:** Our data indicate that PCV21 could prevent 1.5–1.7 times as many pneumococcal disease cases as PCV20 among adults aged ≥65 years in Japan. However, PCV20 should be considered if the incidence of adult IPD cases caused by PCV20-non-PCV21 serotypes increases.

## 1. Introduction

*Streptococcus pneumoniae* (*S. pneumoniae*) is an important human pathogen coated with structurally diverse capsular polysaccharides. To date, more than 100 serotypes have been identified based on antigenic differences in their capsular polysaccharides [1]. In Japan, the 7-valent pneumococcal conjugate vaccine (PCV7) was introduced for pediatric routine immunization in April 2013 and subsequently replaced with PCV13 in November 2013. The vaccination rate has remained at over 95% in children. In April 2024, PCV13 was replaced with PCV15 for pediatric routine immunization, which was subsequently replaced with PCV20 in October 2024. Owing to this immunization program, the incidence of patients with IPD aged younger 5 years from 2014 to 2019 declined by 21.3% compared to that from 2011 to 2013 [2].

For adults aged 65 years or older, the 23-valent pneumococcal polysaccharide vaccine (PPSV23) was incorporated in Japan’s routine immunization program in October 2014 as a Category B disease for individuals who had not received PPSV23 [3]. The cost of vaccination in this category was partly covered (approximately 30%). From 2015 to 2018, the program targeted individuals aged 65, 70, 75, 80, 85, 90, 95, and 100 years or older and was subsequently repeated from 2019 to 2023. During this period, vaccination coverage among individuals aged 65 years was approximately 40%, as indicated in official reports [4]. In April 2024, this program was revised to be implemented for people aged 65 years who had not previously received PPSV23 as the routine immunization.

The emergence of IPD caused by non-PCV13 serotypes in many industrialized countries, including the United Kingdom, France, and Japan [5–7], promoted the development of higher-valency PCVs, such as PCV15 and PCV20, for use in adults [8–11]. In Japan, PCV15 and PCV20 were approved for use in adults in September 2022 and August 2024, respectively. Previous cost-effectiveness studies indicated that the replacement of PPSV23 with PCV20 in the national immunization program for adults aged 65 years or older would be highly cost-effective in Japan [12,13]. Subsequently, the Japanese government replaced PPSV23 with PCV20 in the national immunization program for older adults, effective April 2026. Given the indirect protection conferred by pediatric vaccination programs using PCV13, as well as the differences in the incidence of disease-causing serotypes between children and adults, a 21-valent PCV was developed to prevent IPD and pneumonia in adults in countries with mature pediatric immunization programs [14]. PCV21 was approved for older adults and immunocompromised adults in Japan in August 2025 and is currently under evaluation for its potential inclusion in the national immunization program for Japanese older adults.

Therefore, in this study, we aimed to estimate the preventable pneumococcal disease burden by PCV20 and PCV21 under a national immunization of older adults in Japan.

## 2. Methods

### 2.1 Study design

A national surveillance program (National Epidemiological Surveillance of Infectious Diseases: NESID) for IPD has been implemented under the Infectious Diseases Control Law of Japan since April 2013. Enhanced surveillance was implemented for adult IPD cases reported to the NESID by the Adult IPD Study Group (https://www.niid.jihs.go.jp/basicresearch/research_department/bac1/20250626_invasive_infect_survey.pdf) in 10 prefectures in Japan between April 2013 and December 2025 [7]. All patients with IPD aged ≥15 years were enrolled in this study, covering 18.1% of the Japanese population in this age group.

A case of IPD was defined as the isolation of *S. pneumoniae* from a normally sterile site, such as the blood or cerebrospinal fluid (CSF), or detection of *S. pneumoniae*-specific DNA targeting *lytA* by polymerase chain reaction (PCR) amplification [15]. Pneumococcal isolates were sent to the Department of Bacteriology I, Japan Institute of Health Security (JIHS), National Institute of Infectious Diseases (NIID), and serotyped [16,17]. Sociodemographic and clinical information, including the episode of preceding influenza, was collected using a case report form and submitted to the Department of Infectious Disease Surveillance at JIHS, NIID [18]. Preceding influenza was defined as the laboratory confirmation of influenza virus typically using an immunochromatography test for rapid detection of influenza A and B viruses within ten days before the onset of IPD.

### 2.2 Definition of serotype categories

Serotype categories were defined was follows: PCV7 serotype (serotypes 4, 6B, 9V, 14, 18C, 19F, and 23F), PCV13 serotype (PCV7 plus serotypes 1, 3, 5, 6A, 7F, and 19A), PCV20 serotype (PCV13 plus serotypes 8, 10A, 11A/E, 12F, 15B, 22F, and 33F), PCV21 serotype (serotypes 3, 6A, 7F, 8, 9N, 10A, 11A, 12F, 15A, 15B, 15C, 16F, 17F, 19A, 20A, 22F, 23A, 23B, 24F, 31, 33F, and 35B), PPSV23 serotype (PCV20 plus serotype 2, 9N, 17F, and 20, excluding serotype 6A), PCV13-non-PCV7 serotype (serotypes 1, 3, 5, 6A, 7F, and 19A) and PCV20-non-PCV13 serotypes (serotypes 8, 10A, 11A/E, 12F, 15B, 22F, and 33F), as previously described [4,14]. PCV21-non-PCV13 serotypes were additionally defined as the combination of PCV20-non-PCV13 serotypes and PCV21-specific serotypes (15A, 15C, 16F, 23A, 23 B, 24F, 31, and 35B). PCV20-non-PCV21 serotypes were defined as the nine serotypes included in PCV20 but not in PCV21. All serotype categories are presented at the bottom of Fig. 2.

### 2.3 Estimated number of vaccine-preventable cases

According to the results of the Community-Acquired Pneumonia Immunization Trials (CAPiTA) and its post-hoc analyses, the vaccine efficacy of PCV13 against IPD is 75% and 45% against pneumococcal community-acquired pneumonia (pCAP). As the preventive effects against both IPD and pCAP were estimated to persist for at least 5 years [19–21], and the routine immunization for adults aged 65 years has been implemented since April 2024 in Japan, we postulated that the preventable number of IPD or pCAP cases can be calculated in a target population of adults aged 65–69 years over 1 year. We also postulated that the preventive effects by PCV20 or PCV21 are equivalent to those by PCV13.

The following data were used: 1) the population aged 65–69 years in Japan in 2025 (7.18 million) [22]; 2) the incidence rate of IPD among those aged 65–69 years in 2017–2019 (3.27 cases per 100,000 person-years, data from the present study) or that of pCAP among those aged 65–74 years in 2012 (5.1 cases per 100,000 person-years) [23]; 3) the proportion of vaccine-covered serotypes for patients with IPD aged ≥65 years in 2023–2025 years (PCV20: 50.6%, PCV21: 75.1%, data of present study) or for patients with pCAP aged ≥65 years in 2019–2022, PCV20: 42.8%, PCV21: 72.3%) [24]; 4) vaccine efficacy (75% for IPD, 45% for pCAP) [19], and 5) vaccination coverage rates of 20%, 40%, 60% 80%, and 100%.

### 2.4. Ethical approval

This study was reviewed and approved by the Medical Research Ethics Committee of JIHS, NIID (approval no. 707) and conducted in accordance with the tenets of the Declaration of Helsinki. The need for informed consent was waived, as the data did not contain any patient identifiers and all samples were collected as part of standard patient care.

### 2.5. Statistical analysis

The proportions of clinical characteristics between the two age groups and the incidence of patients with IPD caused by serotypes covered by the two different vaccines were compared using the *χ^2^* test. The Mantel–Haenszel test was used to evaluate trends in the proportions of cases attributable to each vaccine or serotype category across the four isolation periods.

Statistical significance was set at p < 0.05. All statistical analyses were performed using the IBM SPSS Statistics software (version 28.0.1.0; IBM Corp., Armonk, NY, USA).

## 3. Results

### 3.1 Demographic data of adult patients with IPD during the four study periods

The demographic data of 3,351 patients with IPD were compared across the four study periods: before the COVID-19 pandemic (divided into two: 2014–2016 and 2017–2019), during the pandemic (2020–2022), and post-pandemic (2023–2025) in the 15–64-year (28.3%, n=950) and ≥65-year age groups (71.7 %, n=2,401; Table 1). The proportion of patients with a history of PPSV23 vaccination was significantly higher among individuals aged ≥65 years (p < 0.001). However, the PPSV23 or PCV13 vaccination history did not significantly differ in the 15–64-year age group, nor did the PCV13 vaccination history in the ≥65-year age group. The proportion of adult IPD cases preceded by influenza infection significantly differed across the study periods in both the 15–64-year (p < 0.001) and the ≥65-year age groups (p < 0.001). In contrast, the proportion of cases with COVID-19 co-infection did not significantly differ across the study periods in either age group. Notably, the proportion of cases preceded by influenza decreased during the pandemic period (2020– 2022) and increased again during the post-pandemic period (2023–2025) in both groups. The distribution of clinical presentations also significantly differed across the study periods in both the 15–64-year (p = 0.026) and ≥65-year age groups (p < 0.001).

**Table 1.** Clinical characteristics of 3,351 patients with invasive pneumococcal disease (IPD) between the 15–64-year and ≥65-year age groups.

| Study period | Individuals aged 15–64 years,<br>n=950 (28.3%) |  |  |  |  | Individuals aged ≥65 years,<br>n=2,401 (71.1%) |  |  |  |  |
| --- | --- | --- | --- | --- | --- | --- | --- | --- | --- | --- |
| | 2014–16 | 2017–19 | 2020–22 | 2023–25 | $\chi^2$-test | 2014–16 | 2017–19 | 2020–22 | 2023–25 | $\chi^2$-test |
|  | n (%) | n (%) | n (%) | n (%) | p-value | n (%) | n (%) | n (%) | n (%) | p-value |
| n | 223 | 374 | 138 | 215 |  | 488 | 872 | 369 | 672 |  |
| Male | 139 (62.3) | 212 (56.7) | 87 (63.0) | 120 (55.8) | 0.301 | 302 (61.9) | 529 (60.7) | 210 (56.9) | 406 (60.4) | 0.506 |
| Age, median (IQR) | 56 (43–61) | 55 (45–61) | 56 (47–61) | 54 (43–60) | 0.411<sup>a</sup> | 78 (70–85) | 78 (71–85) | 77 (71–86) | 77 (73–85) | 0.172<sup>a</sup> |
| CMC | 127 (57.0) | 191 (51.1) | 68 (49.3) | 108 (50.2) | 0.392 | 271 (55.5) | 507 (58.1) | 224 (60.7) | 431 (64.1) | 0.018 |
| IC | 81 (36.3) | 91 (24.3) | 43 (31.2) | 65 (30.2) | 0.018 | 152 (31.1) | 263 (30.2) | 121 (32.8) | 208 (31.0) | 0.838 |
| Vaccination history |  |  |  |  |  |  |  |  |  |  |
| PPSV23 | 10 (4.5) | 7 (1.9) | 2 (1.4) | 5 (2.3) | 0.186 | 50 (10.2) | 145 (16.6) | 41 (11.1) | 51 (7.6) | <0.001 |
| PCV13 | (0.0) | (0.0) | 1 (0.7) | 1 (0.5) | 0.301 | 3 (0.6) | 7 (0.8) | 4 (1.1) | 4 (0.6) | 0.820 |
| Preceding influenza | 5 (2.2) | 19 (5.1) | 1 (0.7) | 27 (12.6) | <0.001 | 17 (3.5) | 46 (5.3) | 4 (1.1) | 41 (6.1) | <0.001 |
| COVID-19 co-infection | - | - | 3 (2.2) | 4 (1.9) | 0.837<sup>b</sup> | - | - | 15 (4.1) | 25 (3.7) | 0.782<sup>b</sup> |
| Clinical presentation |  |  |  |  | 0.026 |  |  |  |  | <0.001 |
| Bacteremic pneumonia | 104 (46.6) | 175 (46.8) | 52 (37.7) | 126 (58.6) |  | 317 (65.0) | 582 (66.7) | 207 (56.1) | 473 (70.4) |  |
| Meningitis | 50 (22.4) | 77 (20.6) | 32 (23.2) | 30 (14.0) |  | 66 (13.5) | 83 (9.5) | 36 (9.8) | 55 (8.2) |  |
| Bacteremia without any focus | 44 (19.7) | 82 (21.9) | 36 (26.1) | 34 (15.8) |  | 73 (15.0) | 139 (15.9) | 81 (22.0) | 93 (13.8) |  |
| Other IPD | 25 (11.2) | 40 (10.7) | 18 (13.0) | 25 (11.6) |  | 32 (6.6) | 68 (7.8) | 45 (12.2) | 51 (7.6) |  |
| Fatal outcome | 32 (14.3) | 29 (7.8) | 12 (8.7) | 17 (7.9) | 0.042 | 96 (19.7) | 141 (16.2) | 52 (14.1) | 121 (18.0) | 0.135 |

### 3.2 Proportion of vaccine-covered serotypes among adult patients with IPD by age groups and study periods

The proportions of PCV13, PCV20, and PPSV23-covered serotypes decreased during 2014– 2022 but increased during 2023–2025 among patients aged 15-64 years (Fig. 1A, Supplementary Table S1). Over the entire study period, significant decreasing trends were observed for PCV20 (p = 0.012) and PPSV23 (p = 0.048). The proportion of PCV13 (p = 0.006), PCV20 (p < 0.001), and PPSV23 (p < 0.001)-covered serotypes significantly decreased among individuals aged ≥65 years (Fig. 1B). The proportion of PCV21-covered serotype remained unchanged among individuals aged 15–64 years, whereas a significant decreasing trend was observed among individuals aged ≥65 years (p = 0.026).

**Fig. 1.**
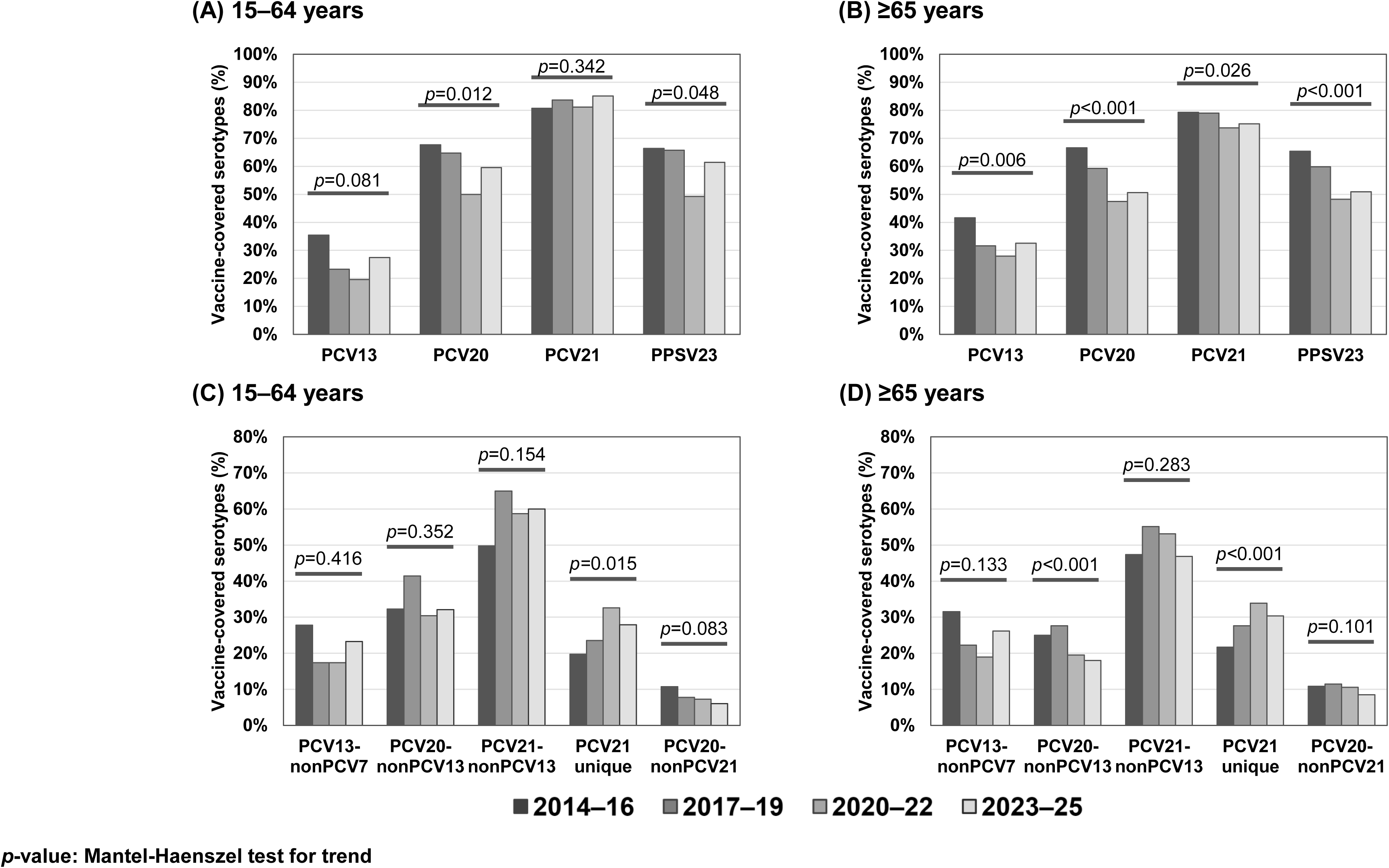
Changes in the proportions of vaccine-covered serotypes causing IPD among adult Japanese patients, stratified by age groups and isolation periods. Serotypes with immunogenicity included in PCV7, PCV13, PCV20, or PPSV23 are presented. p-value, Mantel–Haenszel test for trend in the proportion of each vaccine or each serotype category. Data on the proportions of vaccine-covered serotypes are presented in Supplementary Table S1. PCV7, 7-valent pneumococcal conjugate vaccine; PCV13, 13-valent pneumococcal conjugate vaccine; PCV20, 20-valent pneumococcal conjugate vaccine; PCV21, 21-valent pneumococcal conjugate vaccine; PPSV23, 23-valent pneumococcal polysaccharide vaccine.

Among individuals aged 15–64-years, the proportions of PCV13-non-PCV7, PCV20-non-PCV13, PCV20-non-PCV21-covered serotype remained unchanged, whereas the proportion of PCV21-unique serotypes significantly increased (p = 0.015, Fig. 1C). Among individuals aged ≥65 years, the proportion of PCV20-non-PCV13 (p < 0.001)-covered serotype significantly decreased, whereas that of PCV21-unique serotypes significantly increased during the study period (p < 0.001). In contrast, the proportions of PCV13-non-PCV7, PCV21-non-PCV13 and PCV20-non-PCV21-covered serotype remained unchanged in this age group (Fig. 1D). During 2023–2025, the proportions of PCV20-non-PCV21-covered serotype among patients aged 15–64 years and ≥65 years were 6.0% and 8.5%, respectively.

### 3.3 Distribution of serotypes among adult patients with IPD by age groups and isolation periods

Among individuals aged ≥65 years, the proportion of serotype 3 remained higher than 10%, and that of serotype 19A also remained high. Moreover, the proportions of serotypes 15A and 23A were higher than 5%, and the proportion of serotype 35B significantly increased in this age group (p < 0.05; Fig. 2B). During 2023–2025, the dominant serotypes in PCV20-non-PCV21 among individuals aged ≥65 years were 6B (3%) and 19F (2%).

**Fig. 2.**
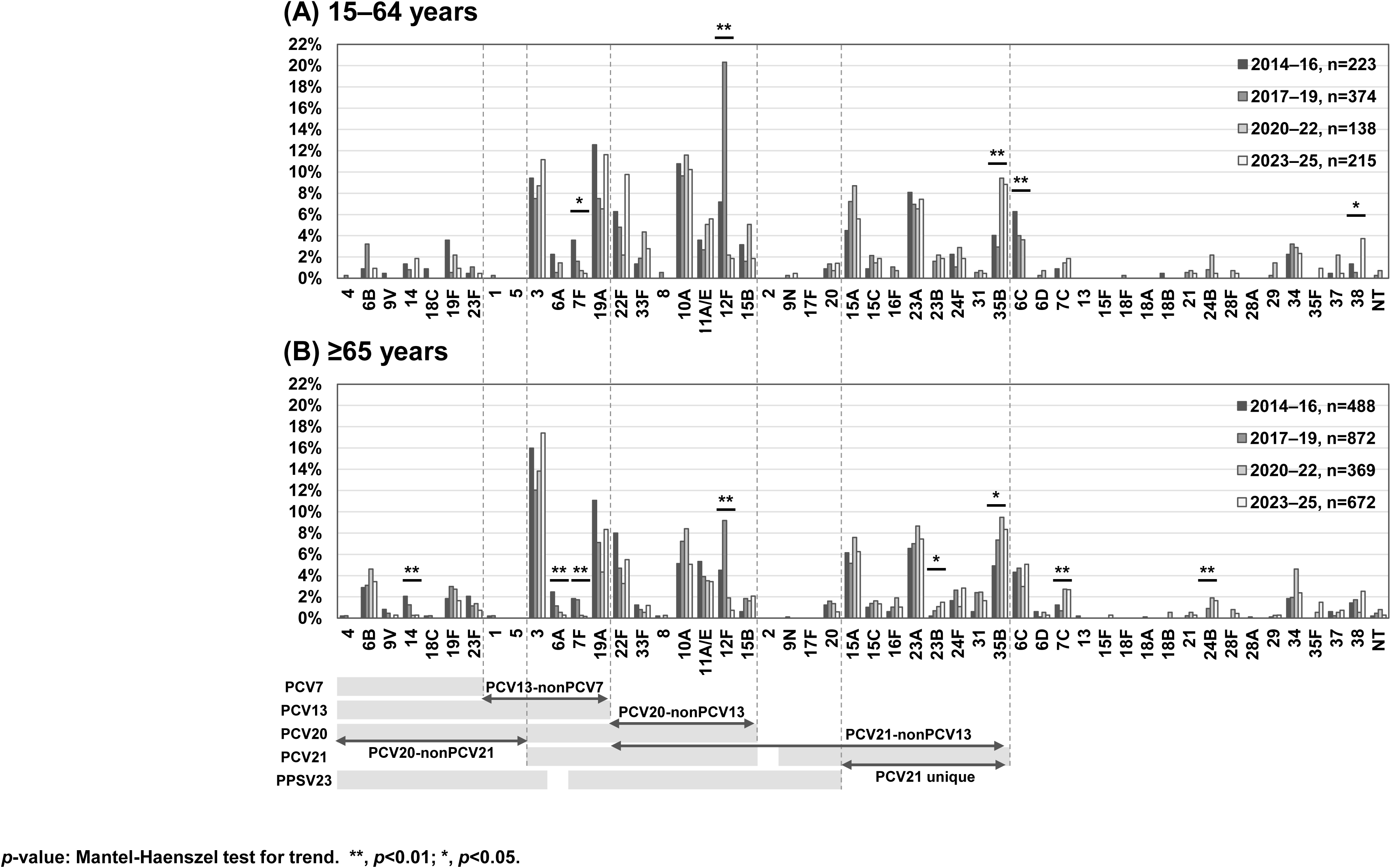
Changes in the proportions of each serotype among adult patients with IPD, stratified by age groups and isolation periods, and serotype coverage of each pneumococcal vaccine. p-value, Mantel–Haenszel test for trend in the proportion of each serotype. NT, nonvaccine type.

### 3.4 Incidence of adult IPD caused by vaccine-covered serotypes during the study periods

The mean annual incidence (cases per 100,000 people) of IPD cases caused by all serotypes among individuals aged 15–64 years was 0.94 before the COVID-19 pandemic, declined by 63% (0.35) during 2020–2022, and slightly increased during 2023–2025 (Fig 3A, 3C). The mean annual incidence of IPD cases among individuals aged ≥65 years was 4.29 pre-pandemic (2017−2019), declined by 59% (1.78) during 2020–2022, and increased up to 89% (3.81) of that pre-pandemic in 2025 (Fig. 3B, 3D). The mean annual incidence of IPD cases caused by PCV21 serotypes was significantly higher than that caused by PCV13 or PCV20 serotypes in the ≥65-year age group (p < 0.001, Fig. 3B). The mean annual incidence of IPD caused by PCV21-non-PCV13 serotypes was also significantly higher than that caused by PCV13-non-PCV7 or PCV20-non-PCV13 in the ≥65-year age group (p < 0.001, Fig. 3D).

**Fig. 3.**
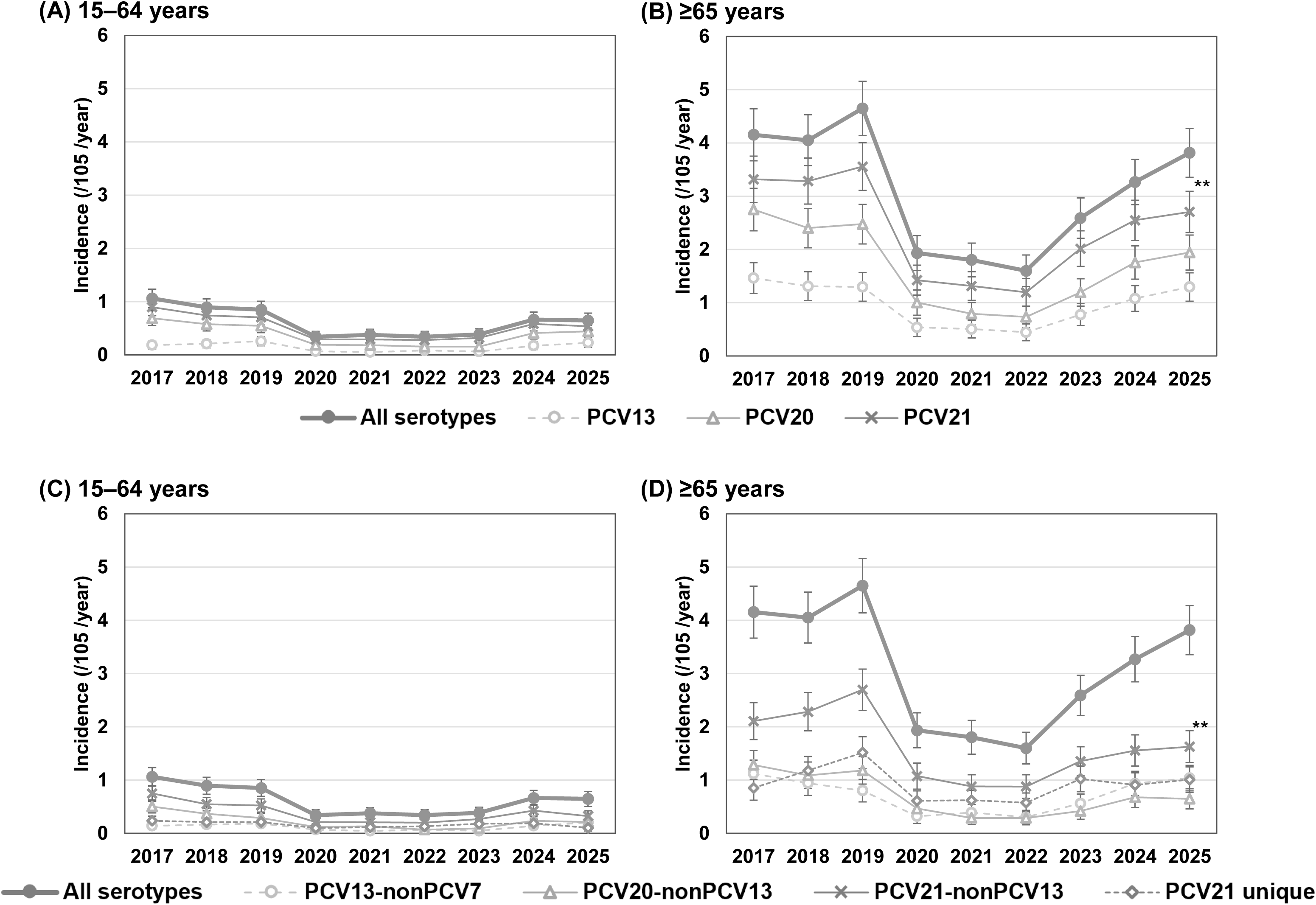
Annual incidences of IPD caused by vaccine-covered serotypes PCV13-nonPCV7, PCV20-nonPCV13, or PCV21-nonPCV13 among adult patients, stratified by age groups and isolation years. PCV21 unique serotypes include 15A, 15C, 16F, 23A, 23B, 24F, 31, and 35B. **p < 0.001, calculated using the *χ^2^* test.

### 3.5 Estimation of the numbers of preventable cases

The numbers of IPD or pCAP cases preventable through routine PCV vaccination among adults were estimated, assuming vaccination coverage rates ranging from 20% to 100% (Table 2). At the current vaccination coverage of 40%, the estimated numbers of preventable cases of IPD by PCV20 and PCV21 were 36 and 53 (person-years), corresponding to reductions of 15.3% and 22.6%, respectively. The numbers of pCAP cases preventable through routine PCV vaccination were similarly estimated under the same vaccination coverage levels. The numbers of pCAP cases preventable by PCV20 and PCV21 were 2,821 and 4,765, respectively, with corresponding reductions of 7.7% and 13.0%, respectively.

**Table 2.** Estimated preventable cases of invasive pneumococcal disease (IPD) or pneumococcal community-acquired pneumonia (pCAP) by vaccination with PCV20 or PCV21.

| PCVs | Vaccination coverages |  |  |  |  |
| --- | --- | --- | --- | --- | --- |
|  | 20% | 40% | 60% | 80% | 100% |
| Estimated preventable cases of IPD (person per year) |  |  |  |  |  |
| PCV20 | 18 | 36 | 54 | 71 | 89 |
| PCV21 | 26 | 53 | 79 | 106 | 132 |
| Estimated reduction of IPD cases (%) |  |  |  |  |  |
| PCV20 | -7.7 | -15.3 | -23.0 | -30.2 | -37.9 |
| PCV21 | -11.1 | -22.6 | -33.6 | -45.1 | -56.2 |
| Estimated preventable cases of pCAP (person per year) |  |  |  |  |  |
| PCV20 | 1,411 | 2,821 | 4,232 | 5,642 | 7,053 |
| PCV21 | 2,383 | 4,765 | 7,148 | 9,531 | 11,914 |
| Estimated reduction of pCAP cases (%) |  |  |  |  |  |
| PCV20 | -3.9 | -7.7 | -11.6 | -15.4 | -19.3 |
| PCV21 | -6.5 | -13.0 | -19.5 | -26.0 | -32.5 |

PCV7, 7-valent pneumococcal conjugate vaccine; PCV13, 13-valent pneumococcal conjugate vaccine; PCV20, 20-valent pneumococcal conjugate vaccine; PCV21, 21-valent pneumococcal conjugate vaccine; PPSV23, 23-valent pneumococcal polysaccharide vaccine.

## 4. Discussion

In the present study, we demonstrated that the mean annual incidence of IPD caused by all serotypes among individuals aged ≥65 years had returned to 89% of the pre-COVID-19 pandemic level in 2025, during the post-pandemic period in Japan. Resurgence of IPD following the COVID-19 pandemic has been reported in several countries, including Switzerland, Germany, and Israel in 2021 [25–27], and in Canada, Italy, and Spain in 2022– 2023 [28–30]. In Japan, the resurgence of IPD following the pandemic was slower than that observed in other countries. Although the reasons for this slower resurgence remain unclear, it may be, in part, attributable to a slow process of relaxation of non-pharmaceutical interventions (NPIs), including mask wearing, and an absence of influenza epidemic in older adults.

In terms of NPIs, a previous study from Germany indicated that decreases in mobility related to transportation, workplaces, and retail activities were correlated with decreases in IPD incidence, and increases in the overall stringency of NPIs were correlated with decreased IPD incidence [26]. A recent discussion paper from Japan on a mask survey reported that mask use remained high among Japanese citizens until mid-March 2023, after which mask use declined; however, the discontinuation of mask use was uneven across the population [31]. Another internet-based survey of 33,000 Japanese participants conducted in 2023 examined the association between mask-wearing behavior and risk factors for severe COVID-19 and indicated that older age, no history of COVID-19, and an increased number of comorbid conditions were associated with increased rates of mask wearing [32].

In terms of influenza activity, we identified an association between preceding influenza infection and an increase in IPD cases following the COVID-19 pandemic, which is consistent with our previous findings from the pre-pandemic period [18]. Therefore, we further evaluated the relationship between national sentinel surveillance data for influenza and the monthly number of adult IPD cases (Supplementary Fig. 1A-D) [33]. From March 2020 to December 2022, we observed no increase in the number of influenza cases among children (<15 years) or adults (15–64 years and ≥65 years) across all 47 prefectures in Japan. In contrast, the monthly number of adult IPD cases during 2020–2022 was lower than that during 2017–2019 or 2023–2025. These findings suggest that the absence of an influenza epidemic was associated with a decreased incidence of IPD among adults during the pandemic. In addition, we noted that in 2023, the influenza epidemic primarily affected children and young adults, but not older adults. In contrast, sharp peaks in reported influenza cases among children and adults (15–59 years and ≥60 years) were associated with an increase in adult IPD cases, primarily among those aged ≥65 years. These findings suggest that a preceding influenza infection among older adults may contribute to increases in IPD incidence in this age group. Therefore, our data indicate that the absence of an influenza epidemic in 2020–2022 and low influenza activity among older adults in 2020–2023 may have contributed to the delayed resurgence of IPD following the COVID-19 pandemic.

Overall, these results suggest that the delayed resurgence of adult IPD in Japan may be attributable to slow relaxation of NPIs and an absence of influenza epidemic among older adults, which is closely related to the implementation of NPIs [26].

In the present study, the incidence of IPD caused by PCV21-non-PCV13 serotypes was significantly higher than that caused by PCV13-non-PCV7 or PCV20-non-PCV13 serotypes among individuals aged ≥65 years between 2017 and 2025. This observation indicates that PCV21 may prevent more IPD cases than PCV20 in this age group. Therefore, PCV21 may be considered more advantageous than PCV20, which is currently used for routine immunization among adults aged 65 years in Japan. A recent study indicated that the serotype coverage of PCV21 among IPD cases was 16–33% higher than that of PCV20 among adults aged 65 years in the United States, Canada, United Kingdom, France, Spain, and Australia [34]. The authors concluded that PCV21 could potentially prevent more IPD cases than PCV20 among patients aged ≥65 years, which is consistent with our findings.

Although we estimated the numbers of IPD or pCAP cases preventable by vaccination with PCV20 or PCV21 among adults aged 65 years at the current vaccination coverage rate of 40% in Japan, the estimated numbers of preventable cases may be insufficient to achieve a substantial impact through routine immunization of older adults. If vaccination coverage increased from 40% to 60%, the estimated number of preventable IPD or pCAP cases would increase to approximately 50 or 4,200 by PCV20 and approximately 80 or 7,100 by PCV21, respectively (Table 2). Therefore, increasing vaccination coverage is important and could significantly enhance the impact of routine immunization among older adults, as previously reported [35]. We also found that the estimated number of preventable cases by PCV21 was 1.5 –1.7 times higher than that by PCV20 for both diseases. Moreover, the estimated number of preventable pCAP cases was approximately 80–90 times higher than that of preventable IPD cases. Although the proportion of PCV21-non-PCV13 remained unchanged, that of PCV21-unique serotypes significantly increased in both age groups. This finding may be partly explained by the composition of PCV21, which comprises replacement serotypes, such as 15A, 23A, and 35B. In contrast, PCV21 lacks PCV7 serotypes and serotypes 1 and 5. PCV20-non-PCV21, including serotypes 6B and 19F, accounted for 8.5% of all isolates from IPD cases among adults aged ≥65 years during 2023–2025 in this study. Therefore, careful monitoring of IPD cases caused by PCV20-non-PCV21 serotypes will be important following the future introduction of PCV21 for routine immunization among older adults. Notably, the indirect effects of pediatric immunization with PCV20, which was introduced in Japan in October 2024, may reduce, at least in part, the burden of disease caused by PCV20-non-PCV13 serotypes among older adults, as suggested by a recent global study [36].

This study had some limitations. First, selection bias may have occurred between enrolled and non-enrolled IPD cases reported to the NESID; among 4,271 adult IPD cases registered to the NESID during 2013–2024, 1,267 (29.7%) were not included in this study. The number of IPD cases registered to NESID in 2025 was not available at the time of analysis. Second, the estimated number of preventable IPD or pCAP cases may not be accurate, as the incidence and proportions of vaccine-covered serotypes for pCAP cases were derived from different study periods. Third, the number of IPD cases may have been underreported, given the increased burden on the healthcare system inflicted by the COVID-19 pandemic during 2020–2022. Finally, because data on the extent to which routine PCV20 vaccination in children indirectly decreases the incidence of IPD among adults remain unavailable, we did not account for this factor in our estimates.

## 5. Conclusions

Our results suggest that both PCV20 and PCV21 significantly decrease the burden of IPD and pCAP among older adults in Japan, with PCV21 estimated to prevent 1.5–1.7 more cases than PCV20. However, PCV20 should still be considered in case of increased incidence of adult IPD cases caused by PCV20-non-PCV21 serotypes. It is crucial to monitor the serotype-specific incidence of IPD following the introduction of PCV21 into the routine vaccination program among older adults in Japan.

## Acknowledgments

We sincerely thank the staff of the local public health centers who collected bacterial isolates and case report forms from the hospitals and the staff of the public health institutes and laboratories.We also acknowledge the following members: Reiko Matsunaga of Infectious Disease Surveillance, JIHS, NIID; Chikako Tsubata of Niigata University Graduate School of Medical and Dental Sciences; Masayuki Ishida of Chikamori Hospital; Junichiro Nishi and Naoko Imuta of Kagoshima University Graduate School of Medical and Dental Sciences; Atsuhito Ikeda of Department of Health and Welfare, Hokkaido Government, Yumiko Maeda of Sapporo City Institute of Public Health, Mikako Hosoya of Niigata Prefectural Institute of Public Health and Environmental Sciences, Kazuki Sakai of Niigata City Institute of Public Health and Environment, Marino Oichi of Mie Prefecture Health and Environment Research Institute, Masami Uchida of Kochi Prefectural Institute of Public Health and Environment, Wakako Sakata of Kitakyushu City Institute of Health and Environmental Sciences, Eriko Kumamoto of Fukuoka City Institute of Health and Environment, Hiroaki Shigemura of Fukuoka Institute of Health and Environmental Sciences, Aoi Endo of Kagoshima Prefectural Institute for Environmental Research and Public Health, Tsuyoshi Kudeken of Okinawa Prefectural Institute of Health and Environment.

## CrediT author contributions

**Conceptualization:** KT and K Oishi; **Data curation**: BC, RS, KG, YT, K Kuronuma, K Oshima, TM, MN, SA, K Kasahara, and TO; **Formal analysis**: BC, RS, and KT. **Writing-review& edit**: MN, YK. **Writing – original draft**: K Oishi. **Funding acquisition**: YA, K Oishi.

## Data availability

Data will be made available upon reasonable request.

