## Supplementary materials for "Estimation of preventable pneumococcal disease burden by PCV20 or PCV21 under the national immunization program for adults aged 65 years in Japan"

**Supplementary materials.** Supplementary Fig. Monthly number of adult IPD cases in the 15–64-year age group and the ≥65-year age group between 2017 and 2025 (A), and the weekly number of cases of influenza per sentinel site in the < 15-year age group (B), 15–59-year age group (C) and the ≥60-year age group (D) in all 47 prefectures, Japan between 2017 and 2025. Notifications of influenza were obtained from the Infectious Disease Weekly Report, which was sourced from the NESID data [33]*.* Data on influenza cases were collected from approximately 5,000 sentinel sites through epidemiological week 14 of 2025 and from approximately 3,000 sentinel sites from epidemiological week 15 of 2025 onward [33]. IPD; invasive pneumococcal disease. NESID; national epidemiological surveillance of infectious diseases.

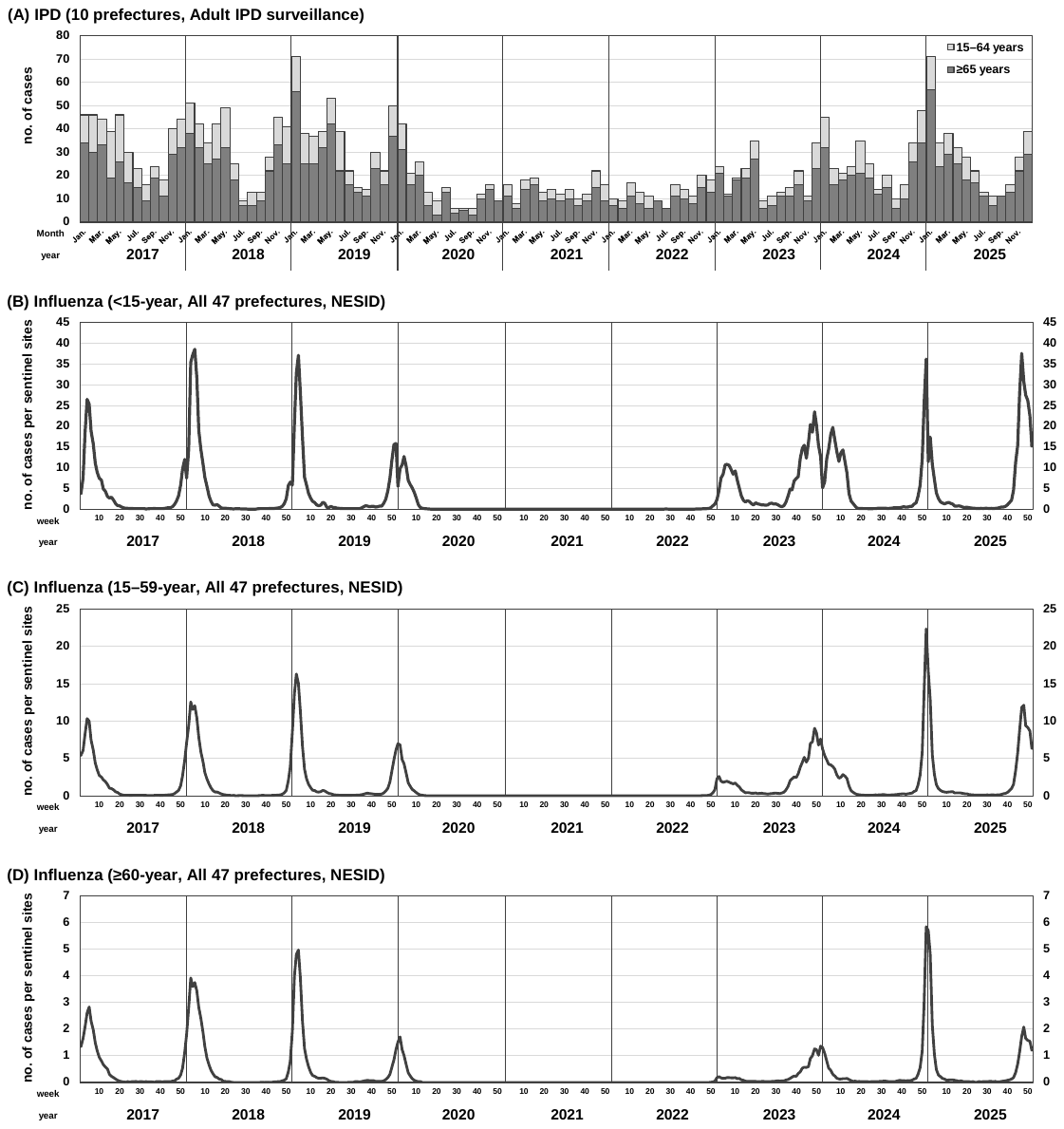

Supplementary Table. Proportions of vaccine-covered serotypes in adult IPD cases, stratified by age-groups and isolation periods.

| Age group | Study period | PCV13 | PCV20 | PCV21 | PPSV23 | PCV13- nonPCV7 | PCV20- nonPCV13 | PCV21- nonPCV13 | PCV21  unique | PCV20- nonPCV21 |
| --- | --- | --- | --- | --- | --- | --- | --- | --- | --- | --- |
| 15–64 years | 2014–16 | 35.4% | 67.7% | 80.7% | 66.4% | 27.8% | 32.3% | 49.8% | 19.7% | 10.8% |
|  | 2017–19 | 23.3% | 64.7% | 83.7% | 65.8% | 17.4% | 41.4% | 65.0% | 23.5% | 7.8% |
|  | 2020–22 | 19.6% | 50.0% | 81.2% | 49.3% | 17.4% | 30.4% | 58.7% | 32.6% | 7.2% |
|  | 2023–25 | 27.4% | 59.5% | 85.1% | 61.4% | 23.3% | 32.1% | 60.0% | 27.9% | 6.0% |
| ≥65 years | 2014–16 | 41.6% | 66.6% | 79.3% | 65.4% | 31.6% | 25.0% | 47.3% | 21.7% | 10.9% |
|  | 2017–19 | 31.7% | 59.3% | 79.0% | 59.9% | 22.2% | 27.6% | 55.2% | 27.6% | 11.5% |
|  | 2020–22 | 27.9% | 47.4% | 73.7% | 48.2% | 19.0% | 19.5% | 53.1% | 33.9% | 10.6% |
|  | 2023–25 | 32.6% | 50.6% | 75.1% | 50.9% | 26.2% | 18.0% | 46.9% | 30.4% | 8.5% |

IPD; invasive pneumococcal disease, PCV; pneumococcal conjugate vaccine.
